# Biallelic protein truncating *EXOSC6* variants cause a neurodevelopmental disorder with cerebellar atrophy, ataxia, and global developmental delay

**DOI:** 10.64898/2026.07.30.26359120

**Authors:** Khondakar Sayef Ahammed, Renzo Guerrini, Milo B. Fasken, Anita H. Corbett, Ambroi van Hoof, Davide Mei

## Abstract

The multisubunit RNA exosome complex provides an essential, highly conserved multifunctional 3’ to 5’ exoribonuclease activity in the eukaryotic nucleus and cytoplasm. Inherited bi-allelic single amino acid variants in the core of the RNA exosome complex have been implicated in causing Mendelian syndromes that affect brain development, collectively termed exosomopathies. The core RNA exosome consists of nine subunits, and pathogenic variants in eight of them (EXOSC1- 5 and EXOSC7-9) have been described in exosomopathies. Here, we describe a patient with cerebellar atrophy, ataxia, and global developmental delay. Trio exome sequencing identified compound heterozygous variants in the final subunit EXOSC6. Previous patients with exosomopathies all have an RNA exosome with only a single amino acid changed, but our patient is missing multiple amino acid residues. The maternal allele is an in-frame deletion that removes 4 amino acids, while the paternal allele introduces a stop codon that removes the last 16 amino acids of EXOSC6. Functional analyses of the variants in a yeast model suggest that both variants are damaging and may affect protein stability. The paternal variant affects a C-terminal α-helix. We tested several other alleles in this helix in our yeast model and show it is important. Overall, our findings broaden the variants implicated in exosomopathies.

## Introduction

Over the last 14 years, pathogenic variants in all but one of the RNA exosome structural genes have been identified and are predominantly linked to neurodevelopmental disease. The RNA exosome is an essential, evolutionarily conserved complex that processes or degrades numerous classes of RNA (1–5). The barrel-shaped, nine-subunit core complex is composed of three cap subunits (EXOSC1-3 in humans) and six PH-like ring subunits (EXOSDC4-9; Figure 1D) (6–9). This core complex can associate with different exoribonuclease subunits, DIS3, DIS3L, and EXOSC10, in different cellular compartments. Key RNA substrates of the RNA exosome include many types of non-coding RNA, including rRNA, snoRNA, snRNA, PROMPTS and enhancer RNAs, as well as aberrant mRNA (1, 10–15). Which specific RNAs are processed by the RNA exosome depends in large part on the association of additional cofactors, or specificity factors (e.g. SKIV2L, SKIV2L2, and HBS1Lv3) with the core exosome.

**Figure 1:**
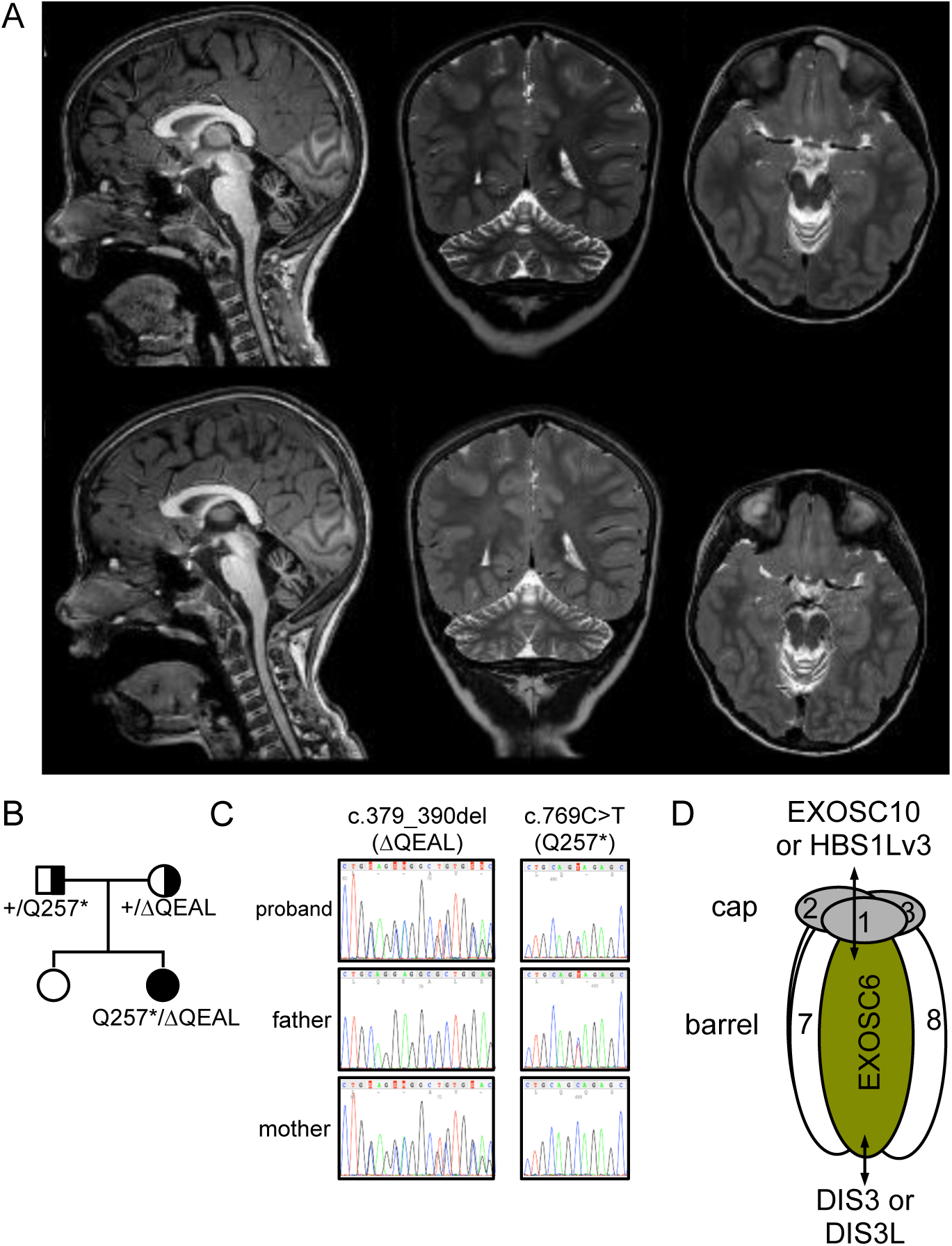
(A) Magnetic resonance imaging (MRI) of Sagittal T1 sections (left panel); Coronal T2 sections (middle panel), and Axial T2 sections (right panel) of the proband’s brain. The top panels represent the MRI obtained when the patient was *[redacted for medrxiv]* and the bottom panels represent the MRI 2 years later. Radiological findings identified cerebellar atrophy involving both hemispheres and the vermis. Pons, brainstem, and mesencephalon do not show signs of atrophy. No progression was observed over the two-year period. (B) Pedigree of the patient with compound heterozygous pathogenic variants in *EXOSC6:* paternally inherited p.(Gln257*) stop-gain variant and maternally inherited p.(Gln127_Leu130del) in-frame deletion variant stop-gain and a maternal in-frame deletion. (C) Sanger sequencing chromatogram of the proband and parents confirming the variants. (D) Schematic of the 9-subunit core RNA exosome complex. EXOSC6 subunit (olive) is one of the 6-barrel subunits that associate with the three- subunit cap. The RNA exosome core binds to the nucleases DIS3 and DIS3L at the bottom of the exosome and EXOSC10 or HBS1Lv3 to the nuclear exosome.

Missense variants in *EXOSC1*, *EXOSC3*, *EXOSC8*, and *EXOSC9* cause pontocerebellar hypoplasia type 1b (PCH1b), missense variants in *EXOSC4* cause developmental delay and intracerebral/basal ganglia calcifications, and missense variants in *EXOSC5* cause developmental delay and cerebellar hypoplasia (16–21). In contrast, missense variants in *EXOSC2* appear to cause diverse types of pathology. An initial report described EXOSC2 variants that cause a syndrome characterized by short stature, hearing loss, retinitis pigmentosa and distinctive facies (SHRF) (22–24). In contrast, a more recent report describes a patient with intellectual disability, epilepsy and microcephaly, which more closely resembles the patients with variants in other EXOSC subunits (25). Three patients with a EXOSC7 variant have previously been mentioned in the literature, but no detailed clinical descriptions are available (26).

Patients with these disorders, defined as exosomopathies, are either homozygous for an *EXOSC* missense variant that changes a single amino acid, compound heterozygous for two different missense variants, or compound heterozygous for a missense variant combined with a nonsense or splice site variant. Analysis of these missense variants in patient-derived cells, tissue culture cells or model organisms indicates that many are deleterious. Many studies have exploited the evolutionary conservation of the RNA exosome complex by employing a budding yeast model system to assess the functional consequences of pathogenic amino acid variants. All structural subunits of the RNA exosome are essential for viability in budding yeast, so this system offers an ideal opportunity to assess function. A number of these variants impair cell growth, reduce RNA exosome function, and, in many cases, decrease subunit protein levels (21, 23, 26–29). Presumably, variants that cause more severe changes than a single amino acid change are not compatible with a successful pregnancy and live birth.

While patients with variants in eight of the nine core RNA exosome subunits (i.e. EXOSC1 to 5 and 7-9) and overlapping neurodevelopmental syndromes have been described, recent reports have identified Mendelian or de novo variants in the exosome catalytic subunits DIS3 and EXOSC10 in patients with more diverse diseases. One study reported de novo loss or inactivation of one *EXOSC10* allele in patients with primary microcephaly, anomalies of cortical structures, intellectual disability and global developmental delay highlighting the role of the nuclear exosome cofactor in brain disorders beyond the RNA exosome core subunits (30). Four of the patients from this study had large deletions (3.8 to 12.3 Mbp) that each affect many genes, but all overlap with EXOSC10. The fifth patient described in this study carried a heterozygous *de novo* missense pathogenic variant in EXOSC10. Other studies have identified Mendelian variants in the RNA exosome catalytic subunits DIS3 and EXOSC10 in patients with primary ovarian insufficiency, but normal neurodevelopment into adulthood (31–33). Somatic *DIS3* variants have also been implicated in multiple melanoma and inherited heterozygous *DIS3* variants appear to cause melanoma susceptibility (34, 35). Finally, Mendelian variants in the SKI complex, a cofactor of the cytoplasmic RNA exosome, cause Tricho-hepato-enteric syndrome, which presents in infants as syndromic diarrhea and woolly hair (36, 37). Thus, although we know that defects in the RNA exosome affect human health, we cannot yet explain the full molecular and pathological consequences of these defects. Further characterization of the spectrum of variants and exosomopathies, and how the variants affect RNA exosome function, should provide additional insight into the molecular mechanisms and spectrum of exosomopathies.

Here, we describe a patient with biallelic variants in the *EXOSC6* gene associated with global developmental delay, ataxic syndrome, and cerebellar atrophy. Exome sequencing of the proband and both parents revealed compound heterozygous variants in the *EXOSC6* gene. This patient is the first known with EXOSC6 variants, and the first patient that has more than a single amino acid change in their RNA exosome. Analysis of these variants and additional ones of the EXOSC6 C-terminal helix in a yeast model reveals that they are damaging and lead to reduced subunit protein levels, indicating that they are likely pathogenic. These results suggest that these *EXOSC6* variants impair RNA exosome function and cause neurodevelopmental disease. With this study, pathogenic variants in all core RNA exosome genes, *EXOSC1* to 9, have now been linked to human Mendelian disease.

## Results

**Clinical Report** [some information is redacted in this version at the request of medrxiv. Details are available from the authors upon reasonable request]

The patient is *[redacted for medrxiv]* born to healthy, non-consanguineous Caucasian parents following an uncomplicated pregnancy and delivery. *[redacted for medrxiv]*. The first clinical evaluation was requested at *[redacted for medrxiv]* because of delayed developmental milestones and marked postural instability. Brain MRI demonstrated isolated cerebellar atrophy (Figure 1A), while spinal MRI findings were normal. *[redacted for medrxiv]* A follow-up brain MRI at *[redacted for medrxiv]* age revealed marked cerebellar atrophy involving vermis and hemispheres, with enlargement of the interfolial spaces and dilated fourth ventricle (Figure 1A). Supratentorial structures and brainstem morphology were preserved.

To identify genetic variants that might be associated with the patient’s phenotype, we performed trio exome sequencing from peripheral blood leukocytes, which identified biallelic variants in the *EXOSC6* gene (NM_058219.3); a paternal nonsense variant [c.769C>T; p.(Gln257*)] and a maternal in-frame deletion variant [c.379_390delCAGGAGGCGCTG; p.(Gln127_Leu130del)] (Figure 1B). Both variants were validated by Sanger sequencing (Figure 1C; Supplemental Figure 1). The Gln257* variant is reported in gnomAD (https://gnomad.broadinstitute.org) with a low allele frequency (2*10^-6;^ 3 out of 1533168 alleles; 0 homozygous), while the in-frame deletion is absent from gnomAD (38).

### Effect of EXOSC6 variants on RNA exosome

The EXOSC6 subunit is one of the six PH-ring subunits and structural studies indicate this subunit interacts with the cap subunit EXOSC1 and neighboring ring subunits EXOSC8 and EXOSC7 (Figure 2A) (8, 39, 40). In addition to these interactions with core RNA exosome subunits, EXOSC6 plays a crucial role in accommodating the nuclear EXOSC10 ribonuclease and the cytoplasmic cofactor HBS1Lv3 (7, 39, 41). We analyzed the patient’s variants in the context of these interactions and structural models of the complex. The Gln257 is located in the C-terminal α-helix of EXOSC6. The Gln257stop variant results in a truncated protein lacking the 16 amino acids of this α-helix (Figure 2A and 2B) (8). These 16 amino acids include residues interacting with the EXOSC1 subunit and the EXOSC10 ribonuclease. The other variant, p.(Gln127_Leu130del), deletes four conserved amino acids within the RNase PH domain of EXOSC6. These four amino acids are at the end of an alpha helix and removing them shortens this helix by approximately one turn and removes a contact with the neighboring subunit EXOSC7 (8).

**Figure 2:**
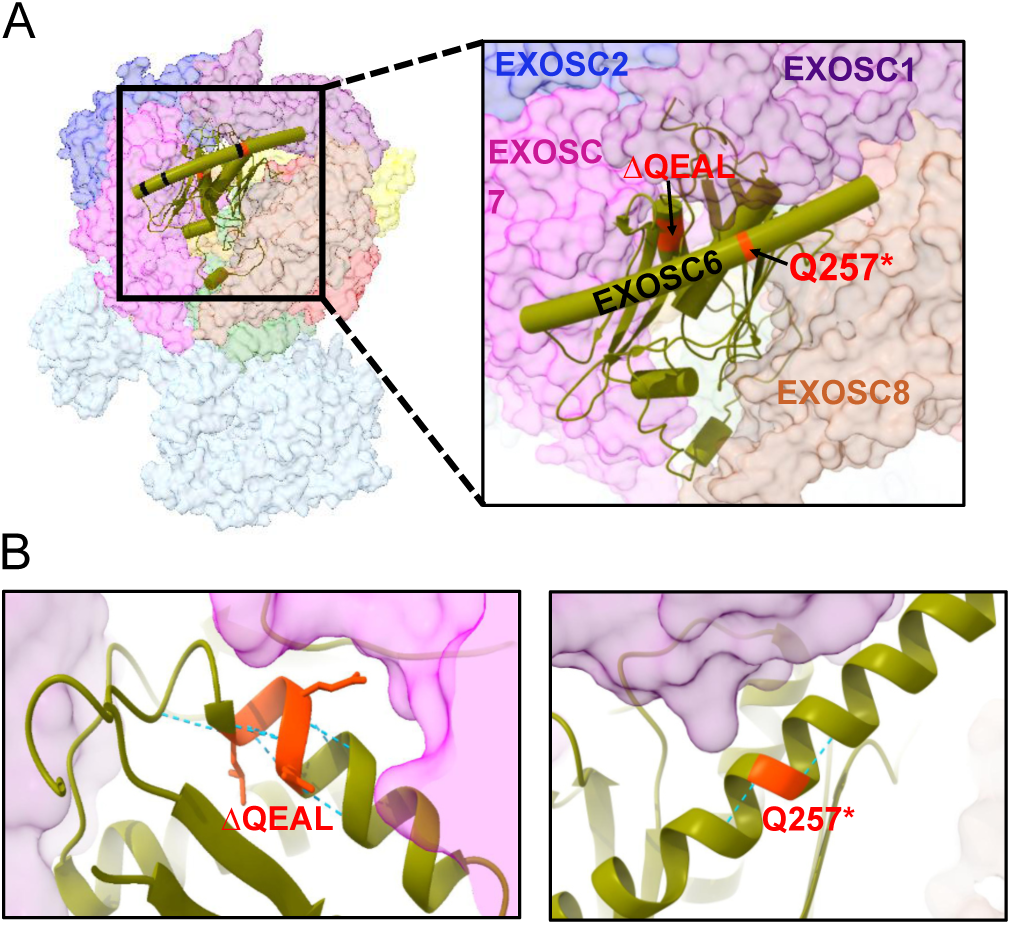
(A) Structure of EXOSC6 (in olive), with amino acids affected in the two variants shown in red in the context of core RNA exosome complex (PDB entry: 6D6R (8)). The neighboring subunits EXOSC1 (purple), EXOSC7 (pink), and EXOSC8 (gray) are shown. (B) Detailed view and side chain of QEAL residues located in the second α-helix (left), and the Q257 residue in the last α-helix of the EXOSC6 (right). The E of QEAL forms one of the contacts with EXOSC7.

### Functional analysis of EXOSC6 variants in a yeast model indicates they are damaging

To determine the effects of EXOSC6 variants, we performed functional analysis using a *Saccharomyces cerevisiae* model. Two different approaches have previously been used to model EXOSC variants in budding yeast. In one approach, the human and yeast proteins are aligned, and the yeast gene is mutated to change the protein with the same (or similar) amino acid change as the human variant. This approach requires that the human and yeast proteins have a similar amino acid at the variant position. For example, EXOSC3-W293R can be modeled in the budding yeast ortholog because the W residues are conserved (23). However, this approach was not readily applicable to EXOSC6 because this subunit has the lowest sequence similarity to the budding yeast ortholog (Figure 3A and Supplemental Figure 2). In particular, the QEAL deleted in the maternal allele corresponds to MGIF in the yeast ortholog. The yeast ortholog has a shorter final alpha helix and therefore the paternal variant could also not be modeled in the yeast ortholog (Figure 3A and Supplemental Figure 2). In a second approach, the yeast gene is completely deleted and replaced by its mammalian ortholog (42). Four of the budding yeast subunits can be successfully replaced by their human ortholog, while two additional ones, including the EXOSC6 ortholog, can be replaced by the mouse ortholog. We thus aligned the highly similar human and mouse EXOSC6 protein sequences. This revealed that human Q257 corresponds to mouse Q258, and that the C-terminal alpha helix of EXOSC6 is of similar sequence and length between human and mouse (Figure 3B and Supplemental Figure 2). The QEAL residues deleted from the maternal allele are also conserved in mouse EXOSC6. We thus introduced corresponding amino acid changes in mouse EXOSC6 (Q258* and ΔQEAL) and tested whether the variants could rescue deletion of the budding yeast ortholog (named *MTR3*). As previously reported, the wild-type mouse EXOSC6 could replace the yeast ortholog. However, the EXOSC6-Q258* and EXOSC6-ΔQEAL alleles failed to complement and do not support growth in the yeast model (Figure 3C). This result indicates that both variants fail to carry out the essential function of the RNA exosome in this yeast model. Based on these functional analyses, the patient-derived variants are damaging to RNA exosome function in this budding yeast model and therefore likely pathogenic.

**Figure 3:**
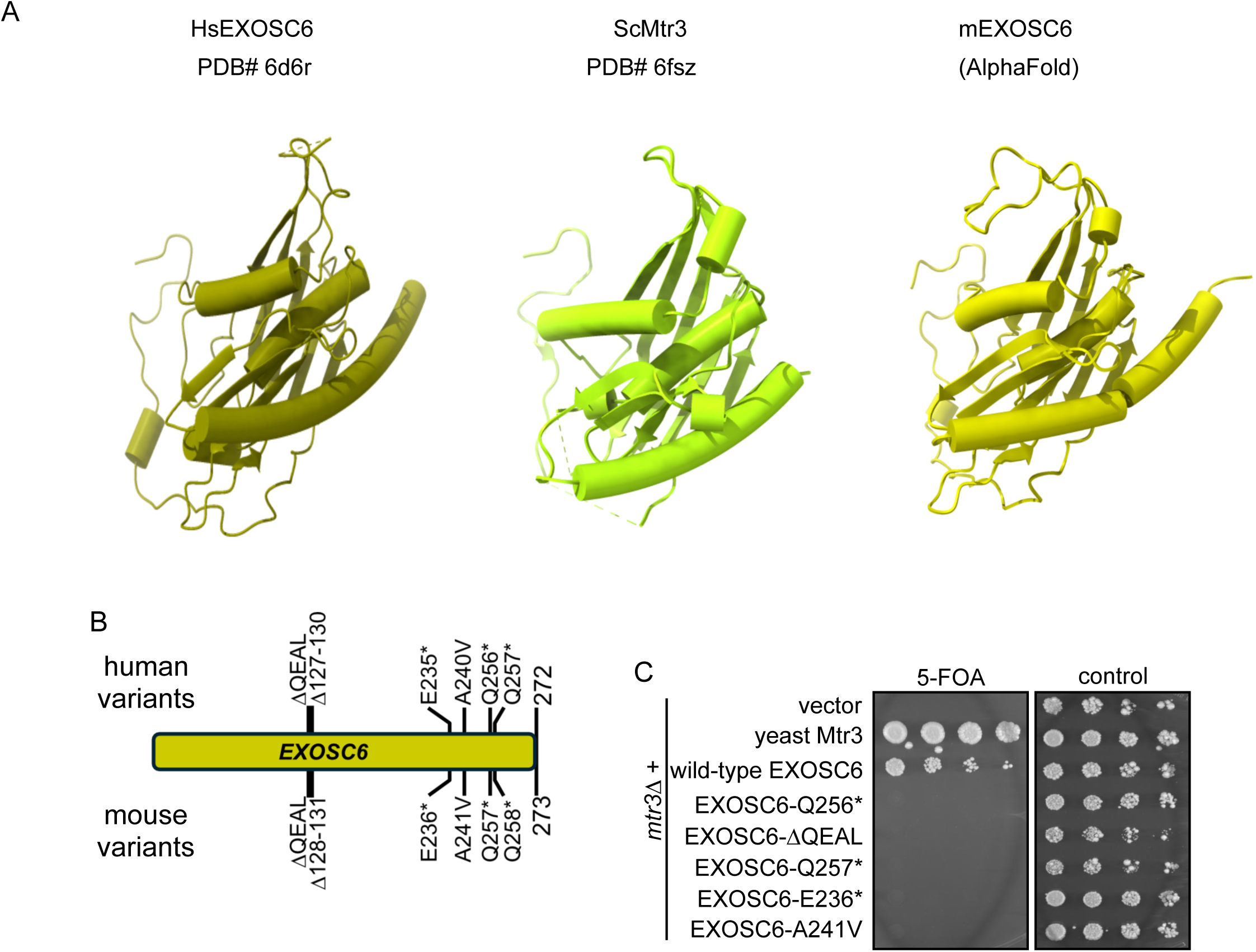
(A) Comparison of human, yeast, and mouse EXOSC6 structures highlighting the overall conservation of the corresponding subunits. The human (olive) and yeast (green) structures are experimentally determined RNA exosome structures (PDB entries: 6D6R (8) and 6FSZ (40)), while the predicted mouse EXOSC6 structure was generated using AlphaFold3 (48). (B) Positions of human EXOSC6 variants and their corresponding mutations in mouse EXOSC6. (C) The effect of EXOSC6 variants was analyzed by assessing the ability of the variants to complement the *mtr3Δ* lethality. The wild-type mEXOSC6 or the variants were expressed in *mtr3Δ* and subjected to plasmid shuffle experiments on media containing 5-FOA or control media lacking leucine and uracil. Serial dilutions were spotted, and growth was recorded after 6 days at 30°C. 5-FOA selects for cells that have lost the *URA3/MTR3* plasmid and growth on 5-FOA therefore indicates that the EXOSC6 allele on the other plasmid is functional.

To determine the effect of the variants on the steady-state protein level of mouse EXOSC6 heterologously expressed in yeast, we compared the levels of wild-type and mutant proteins using an EXOSC6 antibody. Because expression of each of these EXOSC6 variants as the sole copy is lethal in yeast cells, we expressed wild-type and mutant EXOSC6 in yeast cells containing a wild-type copy of the yeast ortholog. The yeast protein is sufficiently different to make it undetectable with this antibody (Figure 4, vector lane). We could readily detect wild-type EXOSC6 expression (Figure 4). However, the EXOSC6-Q258* and EXOSC6-ΔQEAL variants had substantially reduced protein levels compared to wild-type EXOSC6. Notably, the effect of EXOSC6-Q258* appears to be more pronounced and the variant protein was essentially undetectable. The effect of EXOSC6-ΔQEAL on protein level was less than EXOSC6-Q258* (less than 2-fold) but reproducible in three biological replicates (p=0.034 in ANOVA test; Figure 4). The reduced protein level of EXOSC6-Q285* likely explains why this allele failed to complement. We speculate that the slightly reduced level of the EXOSC6-ΔQEAL variant does not fully explain the loss of function of this variant, and that the protein that is produced is not fully functional.

**Figure 4:**
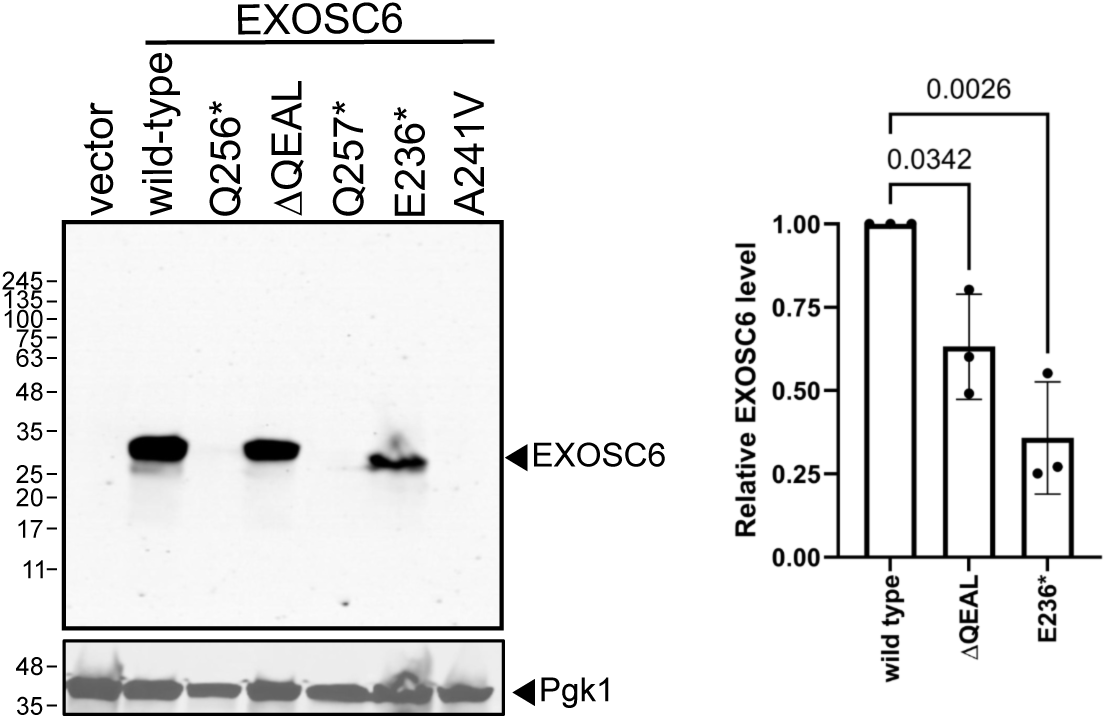
The steady-state protein levels of mEXOSC6 variants were compared to those of the wild-type protein. EXOSC6 was detected using an anti-EXOSC6 antibody raised against the human protein. EXOSC6 levels were normalized to loading control Pgk1 and plotted relative to wild-type expression. Statistical significance was determined using one-way ANOVA.

### Further analysis of the nonconserved C-terminal helix of EXOSC6

As described earlier, the EXOSC6-Q258* variant removes only 16 amino acids from the EXOSC6 protein, and the corresponding alpha helix is shorter in the budding yeast ortholog (Figure 3A and Supplemental Figure 2). We were therefore surprised that shortening this alpha helix in the context of the heterologous expression had such a dramatic effect on protein level and RNA exosome function. During our research we were contacted through GeneMatcher (43) about another patient with EXOSC6 variants. Clinical data on this patient are not available, but both alleles are rare and affect the C-terminal helix: a E235 frameshift variant that completely removes the final alpha helix (gnomAD allele frequency 3.3*10^-6^) and A240V that changes a single amino acid near the beginning of this alpha helix (allele frequency 4.6*10^-6^). To provide additional confidence in our conclusion that small changes to the poorly conserved helix were pathogenic, we analyzed the effect of these variants. We also added the Q256* variant because our patient’s paternal allele changes the second Gln in a Gln-Gln motif. The human Q256* variant that removes one additional residue from the protein and is reported in gnomAD with an allele frequency similar to that of the Q257* seen in our patient (1.3*10^-6^ compared to 1.96*10^-6^). We made all three analogous variants in the mouse gene expressed in yeast (Figures 3 and 4). Expressing mouse EXOSC6-Q257* in budding yeast also fails to complement the yeast deletion. The more dramatic complete deletion of the final alpha helix (EXOSC6-E236*) also failed to complement the yeast deletion, although the protein was present at detectable levels (approximately 3- fold lower than wild type). The more subtle change of a single amino acid in the final helix (EXOSC6-A241V) also failed to complement, and the protein was essentially undetectable. These analyses further indicate that although the C-terminal α-helix of EXOSC6 is naturally shorter in yeast, this helix is critical for RNA exosome function and protein stability.

## Discussion

The clinical-genetic description of the patient with biallelic *EXOSC6* variants included in this report completes the association with exosomopathies of all nine *EXOSC* subunits of the core RNA exosome. The variants in our patient are expected to be more damaging than single amino acid changes. Interestingly, EXOSC6 is among the least conserved subunits of the RNA exosome, indicating that on an evolutionary timescale more variation is permitted in this structural subunit of the complex as compared to the other subunits (26). The final alpha helix of EXOSC6 is naturally shortened in the yeast ortholog. Although this truncation is tolerated in yeast, similar truncation of the alpha helix is damaging in the patient described here and when the mammalian ortholog is expressed in yeast. We hypothesize that deleting this alpha helix in EXOSC6 destabilizes the folding of this protein, but that some other change in the yeast ortholog compensates for this decreased folding stability.

Variants in eight of nine EXOSCs cause neurodevelopmental disorders, although the described symptoms vary. Some variants are also associated with cardiac symptoms(44), which we did not observe in our patient. It is not clear what underlies the phenotypic heterogeneity of exosomopathies. Some symptoms might not be completely penetrant. e.g. RNA exosome defects may increase the probability of cardiac or specific neurodevelopmental defects but not cause these defects in all patients. Each of the variants is extremely rare which eliminates the possibility of linking specific variants to a specific disease presentation. Animal models comparing different variants have promise to extend our understanding of the pathology. Initial studies comparing *EXOSC3* alleles modeled in Drosophila has provided evidence to support a genotype/phenotype correlation (45), but mouse models would provide additional insights. Another possibility is that different RNA exosome subunits have secondary functions that are independent of the RNA exosome complex and defects in these secondary functions contribute to specific disease presentation. Extensive studies on the RNA exosome in mammalian cells appear inconsistent with this second possibility. The complexity of genotype-phenotype correlations is further deepened by some *EXOSC2* variants that cause SHRF instead of neurodevelopmental defects and by variants in the catalytic subunits of the RNA exosome: The EXOSC1-9 core is thought to be non-functional unless the complex associates with either DIS3 or EXOSC10 in the nucleus or DIS3L in the cytoplasm (16, 22, 25). One recent report identified neurodevelopmental defects in patients with *de novo* heterozygous loss-of-function variants of the *EXOSC10* gene (30). This disease process may share a mechanism with the disease process seen in the patient described here and other patients with *EXOSC1-9* variants. Other recent reports have identified Mendelian *DIS3* or *EXOSC10* variants in patients with primary ovarian insufficiency, but normal neurodevelopment into adulthood (31, 32). Somatic *DIS3* pathogenic variants have also been implicated in cancer such as multiple myeloma and other plasma cell dyscrasias (34, 35, 46). Thus, although we know that defects in the RNA exosome affect human health, we cannot yet explain the full molecular and pathological consequences of these defects.

## Materials and Methods

### Identification of EXOSC6 variants by exome sequencing

Genomic DNA from peripheral blood leukocytes of the proband and both parents was extracted using the Qia Symphony SP system (Qiagen, Hilden, Germany) according to the manufacturer’s instructions. Exome sequencing was performed with the Illumina DNA Prep with Exome 2.5 enrichment kit on a NovaSeq 6000DX platform (Illumina, USA), generating 150 bp paired-end reads. The exome sequencing FASTQ data sets were processed for variant calling using the Illumina DRAGEN Germline Analysis Pipeline (Illumina, San Diego, CA). Specifically, we aligned the reads to the GRCh38/hg38 human reference genome and annotated variants with VarSeq (Golden Helix, USA).

Following the analysis, we filtered variants against public (gnomAD v4.1) and in-house databases to retain rare and private variants (MAF <0.1%) located in exons or splice-site regions with potential coding effects. We applied quality filters to include only variants with read depth >10 and genotype quality >20. The variants were evaluated under de novo autosomal-dominant, X-linked, homozygous-recessive, and compound-heterozygous inheritance models. We assessed the functional impact of candidate variants in silico using the dbNSFP database (v3.3a) (47). Finally, we validated the variants and confirmed segregation in the family by Sanger sequencing on both strands using BigDye Terminator v3.1 chemistry and a 3500DX Genetic Analyzer (ThermoFisher, MA, USA). Written informed consent for genetic testing was obtained from the legal guardians of the patient, in accordance with the Declaration of Helsinki and protocols approved by the local institutional review board.

### Structural analysis of the EXOSC6 variants

Structural analysis of the human pathogenic variants was performed using the published cryo-EM structure of the human RNA exosome complex (PDB: 6D6R) (8). The mouse EXOSC variants corresponding to the patient variants were identified by amino acid sequence alignment of the coding sequences of human and mouse EXOSC6. For structural analysis of the amino acid substitutions in the mouse RNA exosome complex, we used AlphaFold to predict the structure of EXOSC6 in complex with the eight other subunits of the core RNA exosome complex (48). All structures presented in this study were analyzed using UCSF ChimeraX.

### Saccharomyces cerevisiae strains and plasmids

The *mtr3* deletion strain (yAv4443: matA, ura3-Δ0, leu2-Δ0, his3-Δ1, lys2-Δ0, mtr3Δ::kanMX [MTR3, URA3]) used in the functional analysis of EXOSC6 variants has previously been described (26). The *MTR3* gene is deleted from its normal chromosomal locus and replaced by an MTR3 rescue plasmid containing the *URA3* counter-selectable marker.

The N-terminal myc-tagged mouse EXOSC6 expression plasmid (pAv2028) under the *TEF1* promoter was described previously (26). EXOSC6 mutant plasmids were generated using QuikChange Lightning Site-Directed Mutagenesis Kit using pAv2028 as template. The mutant plasmid sequences of the mouse EXOSC plasmids were verified by whole plasmid nanopore sequencing (Plasmidsaurus).

### Functional analysis of the variants

For functional analysis of the mouse EXOSC6 variants, the wild-type and mutant plasmids were transformed into the *mtr3* deletion strain (yAv4443), and the transformants were selected on SC media lacking leucine and uracil (SC-Leu-Ura, Sunrise Science). A Plasmid shuffle assay was performed by serially diluting the yeast strains containing empty vector control, wild-type, and mutant mouse EXOSC6 plasmids. The strains were simultaneously spotted onto 5-FOA-containing media to select for cells that had lost the [*MTR3*, *URA3*] plasmid. The growth of the strains expressing mutant mouse EXOSC6 on 5-FOA was compared to the strain that expresses the wild-type mouse EXOSC6 at day 6 at 30°C. Plates with control media were used to ensure that equal numbers of cells were spotted for the different strains.

### Immunoblotting of EXOSC6

The changes in steady-state protein levels of mutant mouse EXOSC6 compared to wild-type mouse EXOSC6 were determined by immunoblotting. Yeast strains expressing either an empty vector, wild-type mEXOSC6, or variants (each containing a wild-type copy of yeast *MTR3*) were grown in SC-Leu,-Ura to an OD600 of ∼0.8 at 30 °C. Cells were then harvested and resuspended in IP50 buffer (50 mM NaCl; 2 mM MgCl₂; 50 mM Tris–HCl, pH 7.5; 0.5 mM β-mercaptoethanol; 0.1% Triton X-100; and 0.1 mM PMSF) supplemented with complete EDTA-free protease inhibitors (Roche). Cells were lysed, and protein extracts were subjected to SDS–PAGE followed by western blot analysis. The Mouse EXOSC6 protein was detected using a rabbit polyclonal primary antibody raised against full length human EXOSC6 (Proteintech; Cat no: 30685-1-AP). As a loading control, yeast Pgk1 protein levels were detected using an anti-Pgk1 antibody (Invitrogen, cat. no. 459250). Secondary detection was performed using goat anti-rabbit (Bio-Rad, cat. no. 1706515) for EXOSC6 and goat anti-mouse (Bio-Rad, cat. no. 1706516) antibodies. Signals were visualized using WesternBright ECL spray (VWR, cat. no. K-12049). Band intensities were quantified and normalized to the Pgk1 loading control. Normalized protein levels of mutant EXOSC6 were compared to wild-type EXOSC6 expression using GraphPad Prism. Statistical significance was assessed by one-way ANOVA.

## Data availability statement

All data relevant to the study are included in the article or uploaded as supplementary information.

## Conflict of interest statement

The authors declare that they have no competing interests

## Ethics statements

### Patient consent for publication

Written informed consent was obtained from all participants or their legal guardians. All procedures complied with the Declaration of Helsinki. Participants gave informed consent to participate in the study before taking part.

### Ethics approval

This study was approved by the Pediatric Ethics Committee of the Tuscany Region as part of the ‘Brain Project - HBOM-Meyer’ (Ref: 283/2021) and the ‘DECODE-EE’ project (Ref: 106/2020).

### AI disclosure statement

No AI tools were used to write this article. The only AI tool used in this project is AlphaFold3, which was used to model the structure on the mouse exosome

## Acknowledgements

We thank the family for their participation in this study. We thank Catherine Stuart for excellent technical assistance and members of the van Hoof lab for insightful comments.

## Funding

This work was funded by the NIH/NIGMS grant R35GM141710 to AvH and Ohio Eminent Scholar funds provided by Ohio State University to AvH. This study was supported, in part, by funds from the ‘Current Research Annual Funding’ of the Italian Ministry of Health. This work was supported by the ‘Brain Project’ by Fondazione Cassa di Risparmio di Firenze (to RG).

## Figure legends

**Supplemental figure 1.**
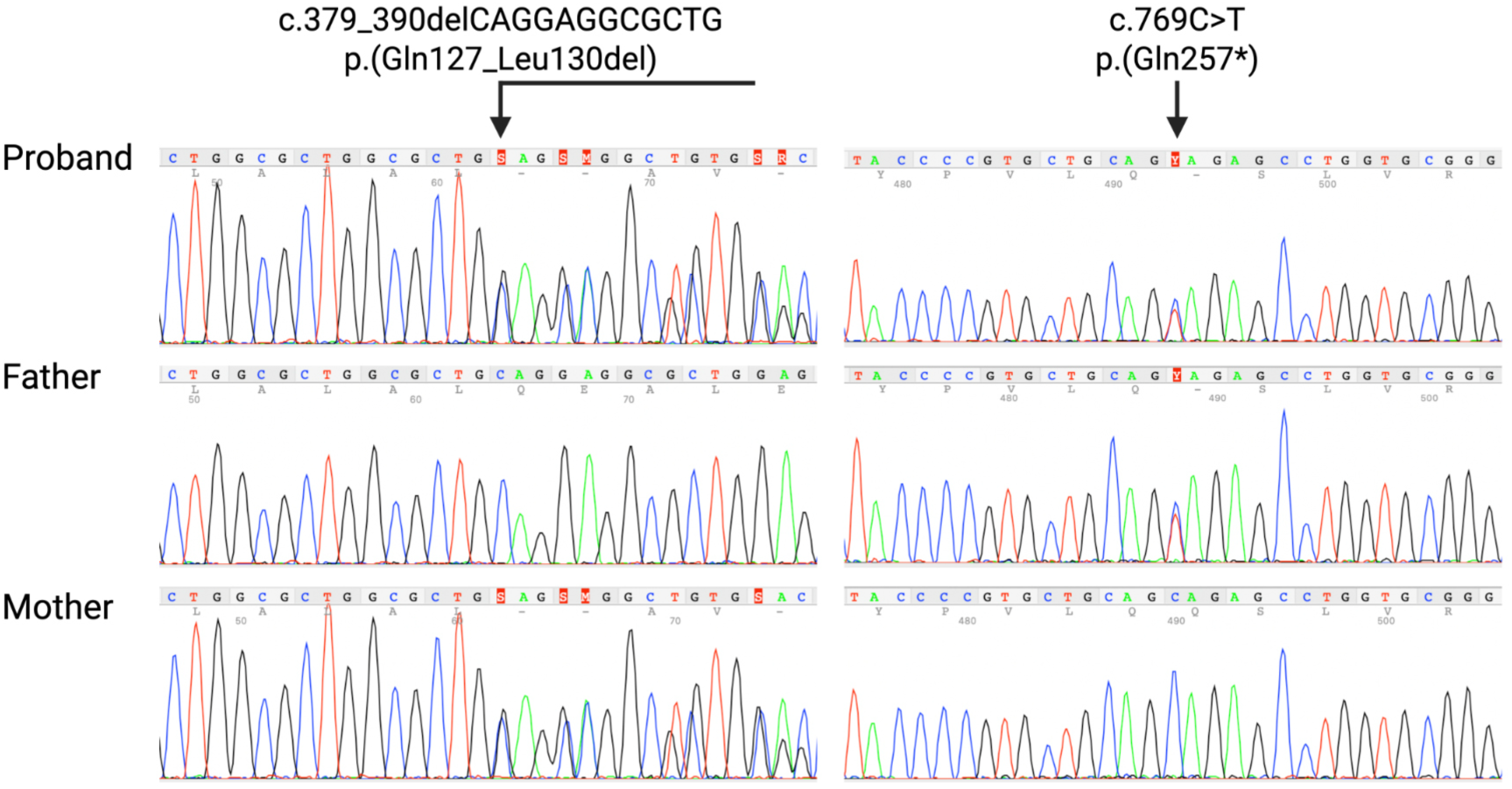
Sanger sequencing chromatograms providing a wider view of the data in figure 1C.

**Supplemental figure 2.**
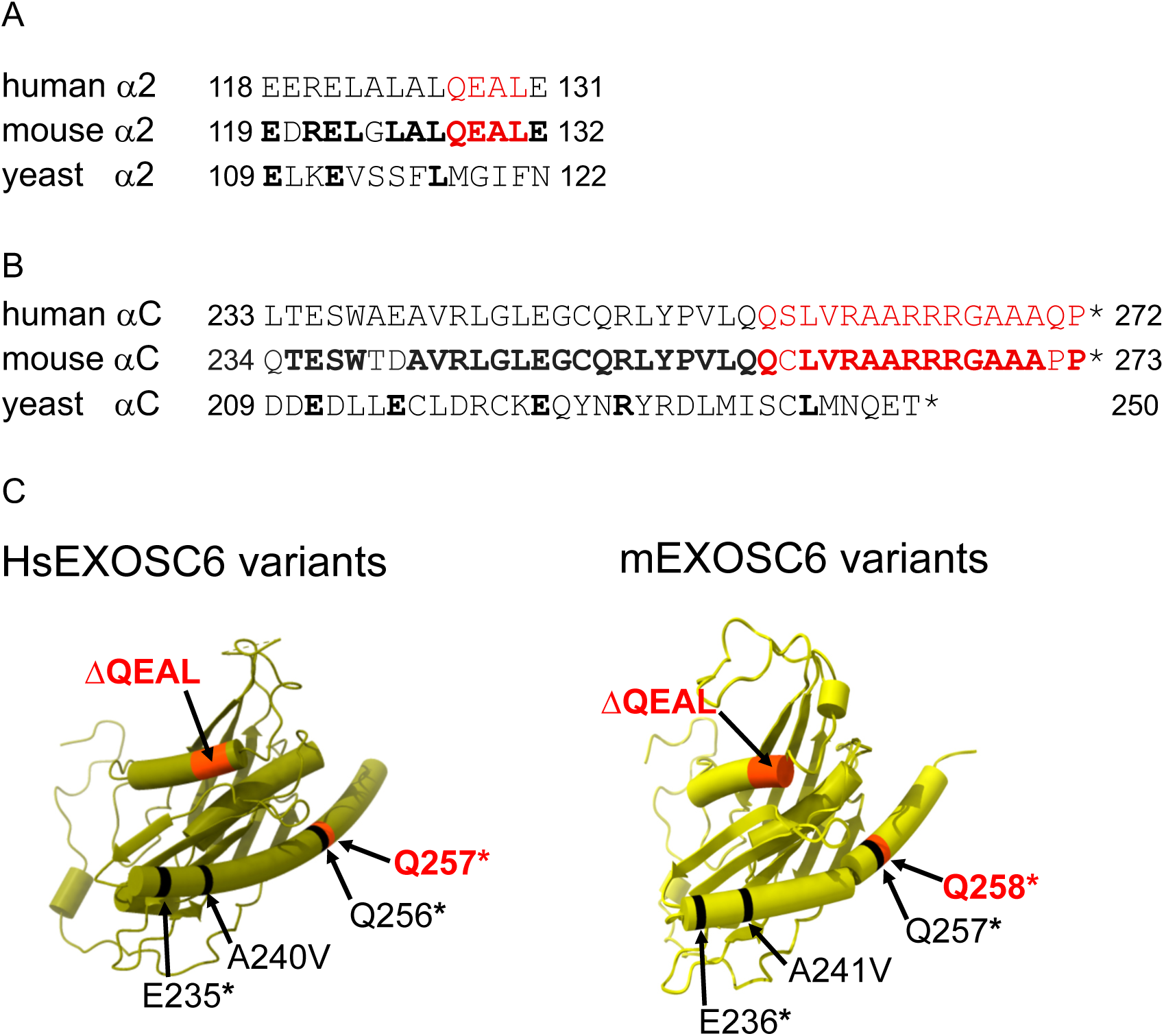
Alignment of second (panel A; α2) and last (panel B; αC) alpha helices of human EXOSC6 with the equivalent mouse and yeast sequences. The human and yeast alignment was generated by aligning the structures (6D6R and 6FSZ) using the matchmaker tool of ChimeraX-1.10.1. The mouse sequence was aligned manually to the human sequence and added to the matchmaker output. Red indicates deleted residues in the human patient and the mouse cDNA expressed in yeast. Bold indicates residues identical with the human sequence. C. The variants are shown on EXOSC6 in the human RNA exosome structure (6D6R; Left) and an AlphaFold model of the mouse RNA exosome.

## Notes

### Competing Interest Statement

The authors have declared no competing interest.

### Author Declarations

This study was approved by the Pediatric Ethics Committee of the Tuscany Region as part of the Brain Project- HBOM-Meyer (Ref: 283/2021) and the 'DECODE-EE' project (Ref: 106/2020).

